# Cognitive screening biases in a secondary prevention Alzheimer’s disease clinical trial

**DOI:** 10.64898/2025.11.28.25341235

**Authors:** Isha Sai, Joshua D. Grill, Kyan Younes, Joseph R. Winer, Karly A. Cody, Reisa Sperling, Elizabeth C. Mormino, Christina B Young

## Abstract

**INTRODUCTION:** Alzheimer’s disease (AD) prevention trials have multiple screening steps to identify cognitively unimpaired individuals with AD biomarker evidence. Cognitive/functional screening tests may be biased in underrepresented groups, thereby impacting trial eligibility.

**METHODS:** 6669 participants screened for the Anti-Amyloid Treatment in Asymptomatic Alzheimer’s (A4) study were grouped by ethnoracial background and testing language. Ethnoracial/language differences in ineligibility reason, cognitive/functional test performance, and amyloid positivity rates were examined.

**RESULTS:** Ethnoracial minorities were least likely to meet eligibility criteria. Patterns of incorrect Mini-Mental State Examination items and impaired Clinical Dementia Rating functional domains differed between groups excluded for impaired cognition/function, suggesting test biases. The Free and Cued Selective Reminding Test yielded more similar exclusion rates across groups than Logical Memory. Cognitive/functional screening biases may impact subsequent biomarker screening as amyloid positivity rates were lowest in ethnoracial minorities.

**DISCUSSION:** Biases in cognitive/functional screening tests may disproportionately exclude ethnoracial minorities in AD prevention trials.

## INTRODUCTION

Clinical trials are necessary for the development of novel therapeutics for Alzheimer’s disease (AD), which currently only has one class of available FDA-approved disease-modifying drugs [1]. Clinical trials for AD are more expensive than trials for many other diseases [2,3] in large part because the time required for participant recruitment typically exceeds the treatment period by 2-3 fold for phase 2 trials and 2-5 fold for phase 3 trials [4]. Recruitment and retention of participants is a major barrier for successful completion of AD clinical trials and there is increasing acknowledgement for the need to determine how potential treatments perform in all populations [5–10]. It is already difficult to recruit participants from diverse backgrounds into a clinical trial for screening, and certain eligibility criteria may unintentionally exclude these participants at higher rates than non-Hispanic White (NHW) counterparts [11–14]. Amongst minoritized populations, Non-Hispanic Asian (NHA) participants are particularly underrepresented in AD trials with factors such as research willingness and inadequate cognitive screening procedures biasing enrollment into biomarker studies [15–17]. Thus, it is critical to carefully examine whether screening eligibility criteria are a contributing factor to the limited inclusion of ethnoracial minorities in AD trials.

The Anti-Amyloid Treatment in Asymptomatic Alzheimer’s (A4) study was a secondary prevention clinical trial that enrolled and randomized cognitively unimpaired (CU) older adults with evidence of elevated brain amyloid to evaluate whether treatment with the monoclonal antibody Solanezumab slowed cognitive decline. Screening procedures for this study were focused on identifying a large cohort of amyloid positive CU individuals to be randomized across treatment and placebo arms. The release of A4 screening data to the scientific community provides a unique opportunity to evaluate the impact of specific inclusion/exclusion criteria on rates of biomarker positivity and trial enrollment. Previous studies examining A4 screening data have shown that ethnoracial minorities were more likely to be excluded after the first screening visit, which involved cognitive testing to establish CU status and clinical assessments to rule out significant medical co-morbidities [14]. Furthermore, lower rates of amyloid positivity have been reported in Non-Hispanic Black (NHB) and NHA individuals compared to NHW individuals [14,18]. These findings show that standard screening criteria may contribute to recruitment bias and limit trial diversity. Although these studies highlight differences in inclusion of ethnoracial minorities in comparison to NHW participants, biases in specific cognitive and functional screening tests, the effect of neuropsychological testing language, and identification of alternative screening measures have not yet been examined.

The A4 study recruited from 67 sites across North America, Australia, and Japan, allowing for the assessment of Japanese vs. English testing language on exclusion rates [19]. Thus, the present study examined the following groups who were screened for the A4 study: NHW participants tested in English (NHW-E), NHB participants tested in English (NHB-E), NHA participants tested in English (NHA-E), NHA participants tested in Japanese (NHA-J), Hispanic White participants tested in English (HW-E), and Other/Multiple races/ethnicities tested in English (O-E). In the context of a secondary prevention clinical trial aimed at enrolling CU older adults with elevated amyloid (A+), our primary aims were to determine whether there are differences between ethnoracial/language groups in reason for exclusion, identify potential biases in the tests used to determine normal cognition and function, assess the performance of alternative cognitive screening measures, and replicate rates of exclusion due to amyloid negativity.

## METHODS

### Participants

All A4 participants provided written informed consent in compliance with local IRBs. Our study included 6669 participants divided into the following ethnoracial/language groups with consideration of the testing language (**Table 1**): NHW-E (n=5739), NHB-E (n=320), NHA-E (n=108), NHA-J (n=156), HW-E (n=170), and O-E (n=176). Our analyses focused on data collected from these participants at all stages of the screening process. Participants who were not tested in their primary language (NHW-E=15, NHB-E=4, NHA-E=16, NHA-J=0, HW-E=21, O-E=8), Hispanic White participants tested in Spanish (n=22), and Other participants tested in Spanish (n=8) were not examined in the present study due to small numbers.

**Table 1.**
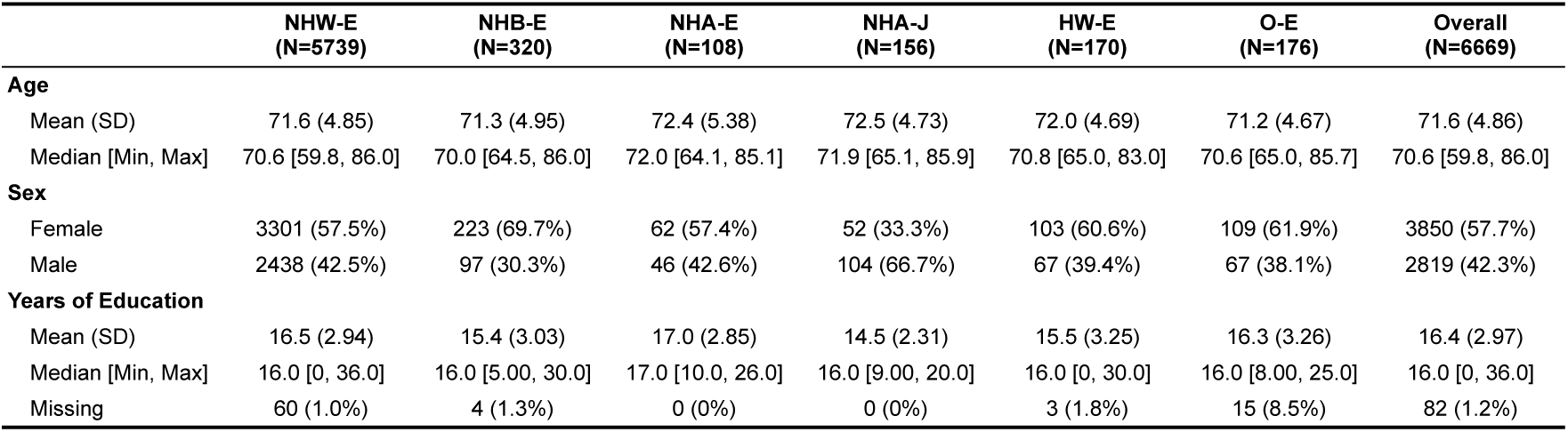
Demographic information for all participants screened for A4 that were tested in their primary language. Note: NHW-E = Non-Hispanic White participants tested in English, NHB-E = Non-Hispanic Black participants tested in English, NHA-E = Non-Hispanic Asian participants tested in English, NHA-J = Non-Hispanic Asian participants tested in Japanese, HW-E = Hispanic White participants tested in English, O-E = Other (Unknown/Multiple Race-Ethnicities) participants tested in English.

|  | NHW-E<br>(N=5739) | NHB-E<br>(N=320) | NHA-E<br>(N=108) | NHA-J<br>(N=156) | HW-E<br>(N=170) | O-E<br>(N=176) | Overall<br>(N=6669) |
| --- | --- | --- | --- | --- | --- | --- | --- |
| **Age** |  |  |  |  |  |  |  |
| Mean (SD) | 71.6 (4.85) | 71.3 (4.95) | 72.4 (5.38) | 72.5 (4.73) | 72.0 (4.69) | 71.2 (4.67) | 71.6 (4.86) |
| Median [Min, Max] | 70.6 [59.8, 86.0] | 70.0 [64.5, 86.0] | 72.0 [64.1, 85.1] | 71.9 [65.1, 85.9] | 70.8 [65.0, 83.0] | 70.6 [65.0, 85.7] | 70.6 [59.8, 86.0] |
| **Sex** |  |  |  |  |  |  |  |
| Female | 3301 (57.5%) | 223 (69.7%) | 62 (57.4%) | 52 (33.3%) | 103 (60.6%) | 109 (61.9%) | 3850 (57.7%) |
| Male | 2438 (42.5%) | 97 (30.3%) | 46 (42.6%) | 104 (66.7%) | 67 (39.4%) | 67 (38.1%) | 2819 (42.3%) |
| **Years of Education** |  |  |  |  |  |  |  |
| Mean (SD) | 16.5 (2.94) | 15.4 (3.03) | 17.0 (2.85) | 14.5 (2.31) | 15.5 (3.25) | 16.3 (3.26) | 16.4 (2.97) |
| Median [Min, Max] | 16.0 [0, 36.0] | 16.0 [5.00, 30.0] | 17.0 [10.0, 26.0] | 16.0 [9.00, 20.0] | 16.0 [0, 30.0] | 16.0 [8.00, 25.0] | 16.0 [0, 36.0] |
| Missing | 60 (1.0%) | 4 (1.3%) | 0 (0%) | 0 (0%) | 3 (1.8%) | 15 (8.5%) | 82 (1.2%) |

### A4 Eligibility Procedures

The A4 study screened 6763 individuals (65-85 years) to identify 1169 A+ CU older adults who were randomized into drug or placebo arms. The screening process involved up to 5 visits over 90 days (**Figure 1**). Participants first underwent initial screening (i.e., Screening Visit 1) in which cognitive and functional assessments were administered to determine cognitive status. Participants with a Logical Memory II (LM) delayed recall score below 6, a Mini Mental State Examination (MMSE) score below 25, and/or a Clinical Dementia Rating (CDR) score greater than 0 were ineligible due to impaired cognition/function (i.e., screening ineligible: impaired cognition/function); participants with a LM score greater than 18 were ineligible due to high cognition (i.e., screening ineligible: high cognition), as these individuals were presumably less likely to be A+. Medical history was also assessed at Screening Visit 1 and those with significant medical comorbidities were ineligible [20]. Whether a participant was ineligible due to a medical or other reason (e.g., voluntary withdrawal from study) was not publicly available, and, as a result, we considered all participants who met cognitive inclusion criteria but did not advance to amyloid PET to be ineligible because of non-cognitive reasons (i.e., screening ineligible: non-cognitive). At Screening Visit 2, participants eligible after cognitive, functional, and medical screening underwent amyloid (^18^F-florbetarpir) positron emission tomography (PET) scanning. Amyloid status was determined using a hybrid quantitative and qualitative method [20]. Those who were A- (SUVR under 1.15, or between 1.10-1.15 without a visual read of confirmed elevated amyloid) were ineligible (i.e., screening ineligible: A-). Participants learned of their amyloid status at Screening Visit 3. A+ participants continued onto Screening Visit 4 for a magnetic resonance imaging (MRI) scan to assess for microhemorrhages and an optional lumbar puncture at Screening Visit 5; additional ineligible participants were identified at this stage (i.e., screening ineligible: post-imaging). This procedure identified 1169 participants who were randomized into drug or placebo arms (**Figure 2**).

**Figure 1.**
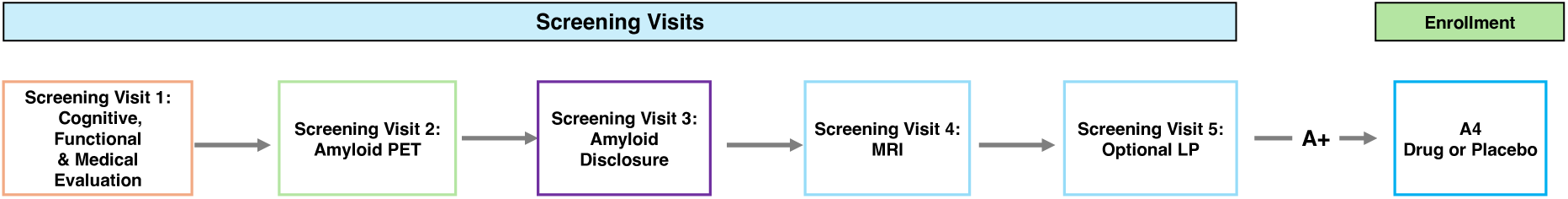
A4 screening procedure to determine trial eligibility across screening visits 1-5. Note: LP = lumbar puncture; PET = positron emission tomography; MRI = magnetic resonance imaging.

**Figure 2.**
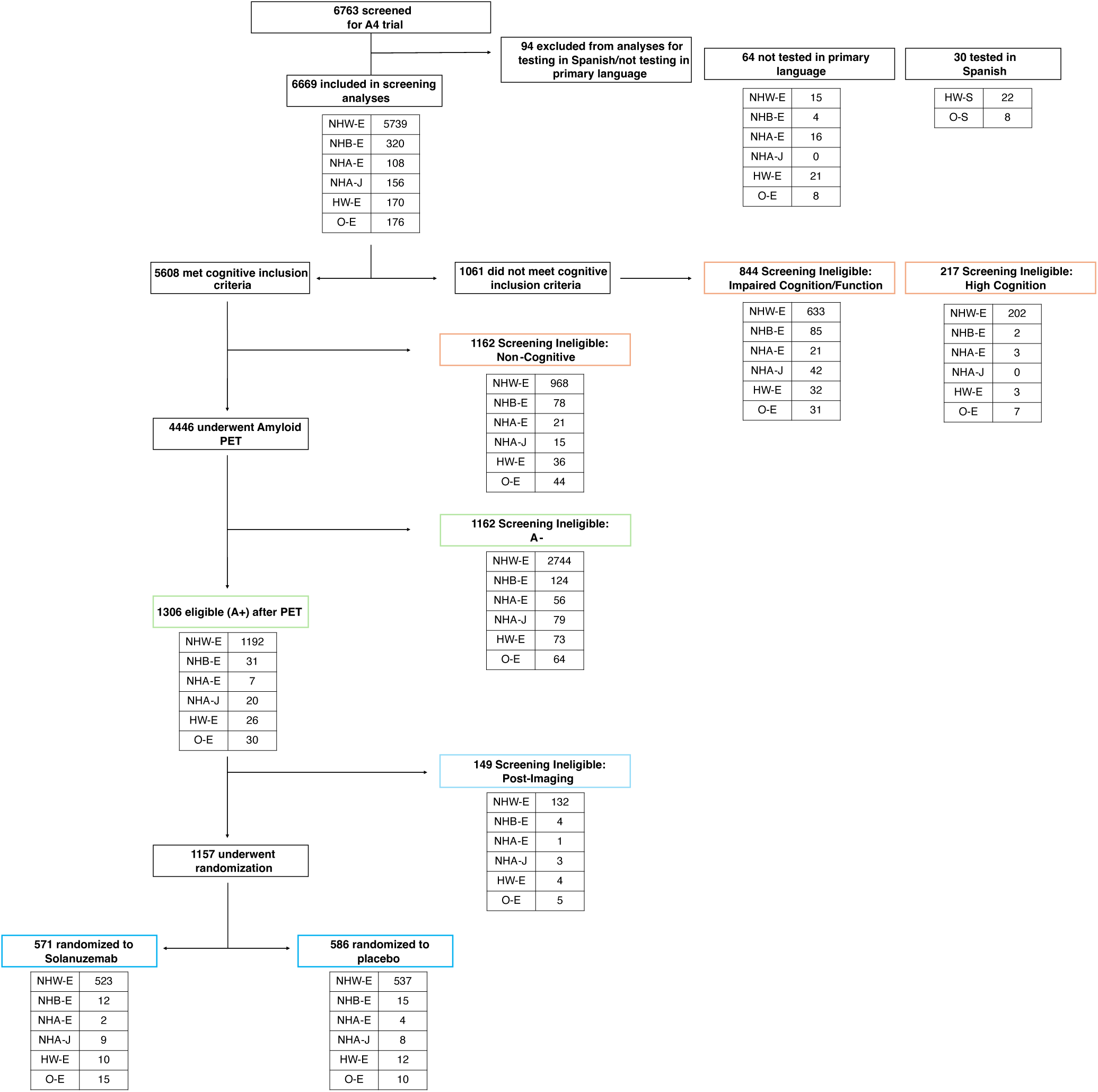
A4 screening and flow chart for all participants. Exclusion reason and number of participants excluded at each stage from each ethnoracial/language group are shown. Note: NHW-E = Non-Hispanic White participants tested in English, NHB-E = Non-Hispanic Black participants tested in English, NHA-E = Non-Hispanic Asian participants tested in English, NHA-J = Non-Hispanic Asian participants tested in Japanese, HW-E = Hispanic White participants tested in English, O-E = Other (Unknown/Multiple Race-Ethnicities) participants tested in English. Colored boxes indicate screening visit number detailed in Figure 1.

### Available Cognitive Data

Item level data were available for the MMSE, a global cognition screening measure, and domain level data were available for the CDR, a measure of functional impairment. Item level data was not available for LM, a test of episodic memory, precluding more detailed examination of incorrect items on this test. In addition to the three screening tests used to determine CU status (i.e., MMSE, LM, and CDR), the Free and Cued Selective Reminding Test (FCSRT) was also used to assess episodic memory but was not used for screening; FCSRT data were available in 6489 participants of the 6669 participants analyzed in this study.

### Statistical Analyses

Statistical analyses were performed using R v4.3.3. First, to evaluate demographic differences between ethnoracial/language groups, an analysis of variance (ANOVA) was used for age and years of education, and Fisher’s exact test was used for sex. Second, we used a series of chi-square tests and logistic regression models to examine whether odds of ineligibility due to impaired cognition/function and non-cognitive reasons differed between ethnoracial/language groups. Third, we examined whether performance on the MMSE and CDR-SOB (CDR Sum of Boxes) differed by ethnoracial/language group using ANOVAs and linear regression models with covariates. Fourth, we compared performance on the FCSRT free recall and LM amongst all screened participants using ANOVAs and linear regression models with main effects of ethnoracial group. We also determined if the FCSRT free recall could be used as a screening instrument that would lead to less bias than LM by examining theoretical exclusion rates if an exclusion criteria of FCSRT free recall < 25 was used instead of LM < 6 [21]; a FCSRT free recall cutoff of 25 corresponds to moderate retrieval impairment in which free recall declines at a constant rate but storage is preserved [21]. Chi-square and logistic regression models were used to determine whether rates of theoretical exclusion based on FCSRT free recall < 25 or LM < 6 differed between ethnoracial/language groups. Fifth, to determine if rates of A+ differed across different ethnoracial/language groups, a chi-square test and logistic regression model were used in all participants who completed amyloid PET. Finally, to determine if rates of exclusion due to post-imaging reasons differed across different ethnoracial/language groups, a chi-square test and logistic regression were used in all A+ participants after amyloid PET. All linear and logistic regression models controlled for age and sex, and analyses involving neuropsychological scores additionally controlled for education.

## RESULTS

### Demographic Differences

6669 participants were screened for the A4 trial and tested in their primary language (**Table 1**). Ethnoracial/language groups significantly differed by age [F(5, 6663)=2.37, p=0.04], sex [p < 0.01], and years of education [F(5, 6581)=27.43, p < 0.001]. The NHA-J group was the oldest group, the only group with more males than females, and the group with the fewest years of education.

### Ethnoracial Minorities are Excluded at Higher Rates due to Cognitive Reasons

Rate of cognitive/functional ineligibility significantly differed between ethnoracial/language groups [χ^2^(5)=62.7, p < 0.001] **(Figure 3).** Notably, 21-27% of ethnoracial minorities were excluded due to high or impaired cognition/function in contrast to 15% of NHW-E participants. Increased odds of exclusion due to high or cognitive/function in all ethnoracial minorities other than HW-E in comparison to NHW-E remained after accounting for age, sex, and education **(Table S1).**

**Figure 3.**
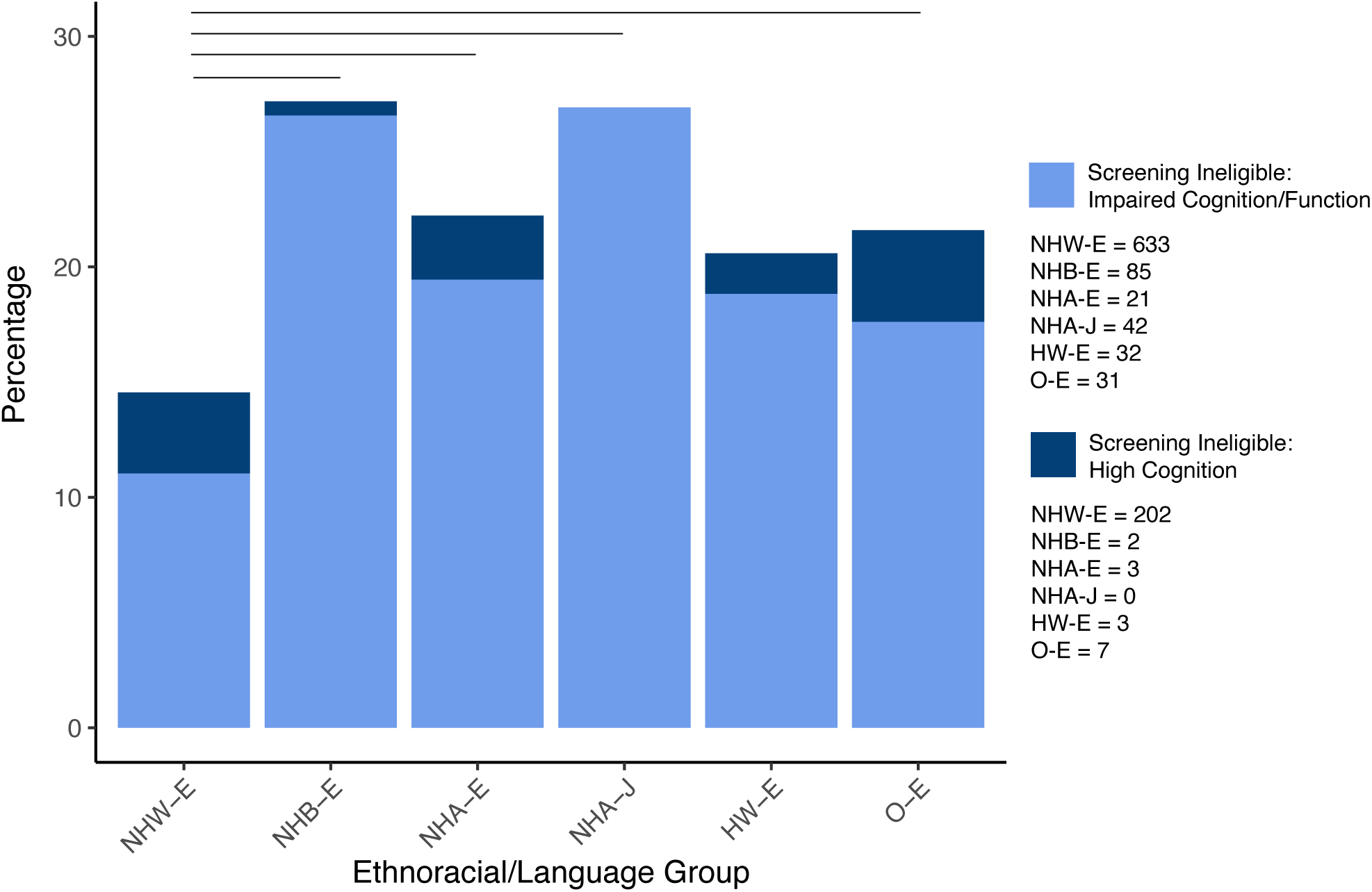
Rates of cognitive/functional screening ineligibility for each ethnoracial/language group. Note: Black bars denote significant differences in percent of cognitive screening ineligible participants using logistic regression adjusting for age, sex, and education. NHW-E = Non-Hispanic White participants tested in English, NHB-E = Non-Hispanic Black participants tested in English, NHA-E = Non-Hispanic Asian participants tested in English, NHA-J = Non-Hispanic Asian participants tested in Japanese, HW-E = Hispanic White participants tested in English, O-E = Other (Unknown/Multiple Race-Ethnicities) participants tested in English

Of all screened participants (n=6669), exclusion due to high rather than impaired cognition/function significantly differed between ethnoracial/language groups [χ^2^(5)=15.1, p=0.001]. Of screened NHW-E participants, 4% were excluded due to high memory performance on LM and 11% were excluded due to impaired cognitive/functional test performance. This is in contrast to 0-4% of ethnoracial minorities who were excluded due to high memory performance on LM and 18-27% who were excluded due to impaired cognitive/functional test performance (**Figure 3**). After accounting for age, sex, and education, the odds of being excluded for impaired cognitive/function test performance remained significantly higher in all ethnoracial minority groups in comparison to NHW-E participants (**Table S2**).

Of participants excluded for high or impaired cognitive/function test performance, the majority of participants were excluded due to performance on only one rather than a combination of measures, although not all participants completed all three screening measures (**Figure 4**). Across ethnoracial/testing language groups, 26-55% were excluded due to low scores on LM only, 26-31% were excluded due to impairments on CDR only, 4-9% were excluded due to low MMSE scores only, and 2-24% were excluded due to high LM scores only. Only 1% of participants had a combination of high cognition (i.e., high LM) and impaired functioning (i.e., high CDR), and there were no participants with a combination of high cognition (i.e., high LM) and low MMSE, providing face validity for these measures.

**Figure 4.**
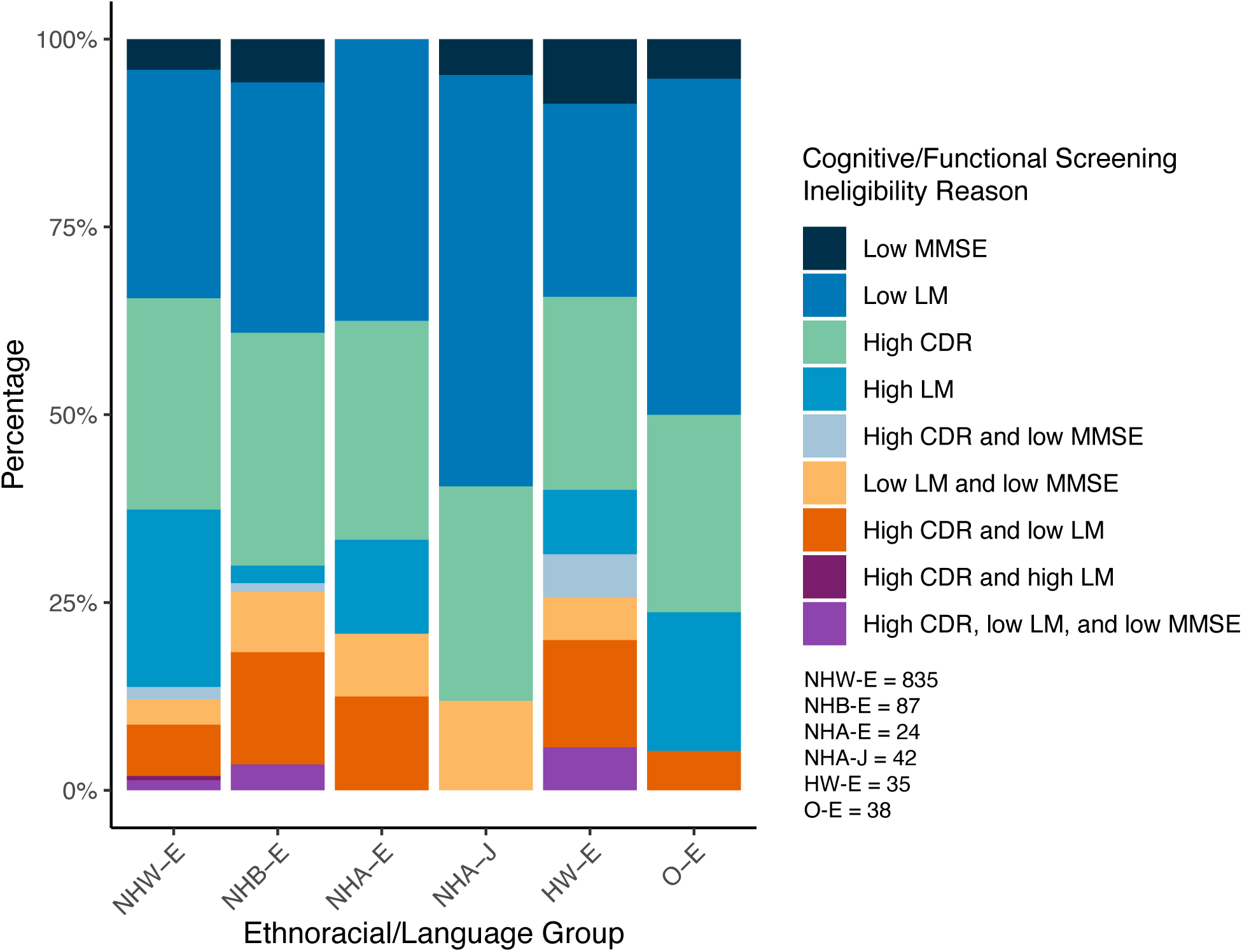
Neuropsychological performance on the three screening tests (MMSE, CDR, and LM) in those excluded due to cognition/function test performance. Note: NHW-E = Non-Hispanic White participants tested in English, NHB-E = Non-Hispanic Black participants tested in English, NHA-E = Non-Hispanic Asian participants tested in English, NHA-J = Non-Hispanic Asian participants tested in Japanese, HW-E = Hispanic White participants tested in English, O-E = Other (Unknown/Multiple Race-Ethnicities) participants tested in English. MMSE = Mini-Mental State Examination. CDR = Clinical Dementia Rating. LM = Logical Memory.

### Performance on Screening Cognitive Tests

When considering all participants screened for A4 (n=6669), ethnoracial/language groups significantly differed on MMSE (**Figure 5A)** [F(5, 6476)=31.5, p < 0.001] with the highest MMSE scores observed in the NHW-E group. Results were similar after accounting for age, sex, and education (**Table S3**). Ethnoracial/language groups also significantly differed in the CDR-SOB (**Figure 5B)** [F(5, 6017)=17.33, p < 0.001]. After accounting for age, sex, and education, CDR-SOB scores were significantly higher for all ethnoracial/language groups other than NHA-J in comparison to the NHW-E group (**Table S3**), indicating higher endorsement of functional impairment in ethnoracial minority groups.

**Figure 5.**
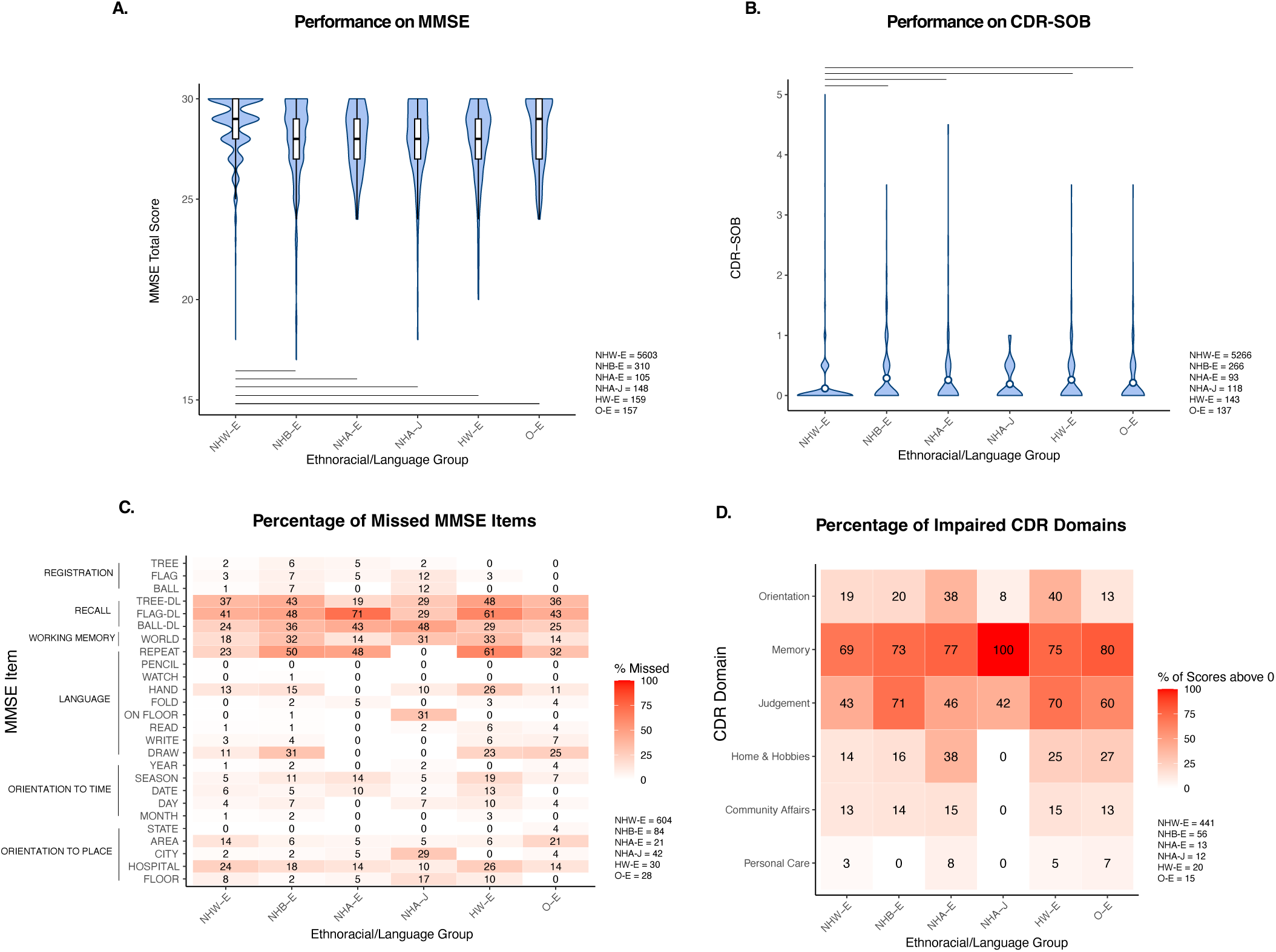
Performance on MMSE and CDR-SOB screening measures as well as item-level patterns for each ethnoracial/language group. Of all screened participants, (A) MMSE scores are highest in the NHW-E group and (B) CDR-SOB scores are lowest in the NHW-E group compared to all other groups. Of participants excluded due to impaired cognition/functioning, (C) patterns of missed MMSE items and (D) reported CDR domains with impairment varies by ethnoracial/language groups. Note: Black bars denote significant differences in MMSE total score and CDR-SOB using linear regression adjusting for age, sex, and education. NHW-E = Non-Hispanic White participants tested in English, NHB-E = Non-Hispanic Black participants tested in English, NHA-E = Non-Hispanic Asian participants tested in English, NHA-J = Non-Hispanic Asian participants tested in Japanese, HW-E = Hispanic White participants tested in English, O-E = Other (Unknown/Multiple Race-Ethnicities) participants tested in English. MMSE = Mini-Mental State Examination. CDR-SOB = Clinical Dementia Rating Sum of Boxes

Given the differing performance on these screening measures by ethnoracial/language group, we next examined domain-specific scores on the MMSE and CDR in participants excluded due to impaired cognition/function (n= 844). Qualitatively, participants had a higher percentage of errors on memory-related items, followed by working memory and sentence repetition items on the MMSE (**Figure 5C**). The qualitative pattern of MMSE errors varied across the ethnoracial/language groups. NHB-E participants had errors on delayed recall memory, working memory, sentence repetition, and visuospatial items at a higher rate, and location orientation items at a lower rate than NHW-E participants. HW-E participants showed a similar pattern with the addition of a higher percentage of errors on time orientation items than the NHW-E participants. NHA-E participants had errors on delayed recall words, sentence repetition, and time orientation items at a higher rate, and the visuospatial item at a lower rate than the NHW-E group. NHA-J participants had a higher percentage of errors during encoding but a lower percentage of errors on 2 out of 3 delayed recall items, as well as a higher rate of errors on receptive language and location orientation items than the NHW-E group. O-E participants had a similar score pattern to the NHW-E group.

Qualitative examinations of CDR domains showed that Memory was the most commonly impaired domain across all ethnoracial/language groups, followed by Judgment, and Orientation (**Figure 5D**). 100% of NHA-J study partners reported impairments in Memory but rarely endorsed other domains. NHB-E, NHA-E, HW-E, and O-E study partners reported impairments in most domains at a similar rate as NHW-E study partners with the following exceptions. In comparison to NHW-E, NHB-E study partners were more likely to report impairments in Judgment; NHA-E study partners were more likely to report impairments in Orientation, Home & Hobbies, and Personal Care; and O-E study partners reported were more likely to report impairments in Memory, Judgement, Home & Hobbies, and Personal Care.

### LM vs. FCSRT Free Recall performance

We next examined LM and FCSRT free recall performance, two episodic memory measures that were and were not used for screening, respectively. When considering all participants screened for A4 (n= 6669), performance on LM significantly differed by ethnoracial/language group [F(5, 6501)=45.78, p < 0.001] with the highest scores observed in the NHW-E group (**Figure 6A**). In contrast, FCSRT free recall performance was only marginally significantly different across ethnoracial/language groups [F(5, 6483)=2.021, p=0.07] in all participants screened for A4 (**Figure 6B**). When adjusting for age, sex, and education, ethnoracial/language differences on LM remained significantly higher in NHW-E than all ethnoracial/language minority groups; group differences on FCSRT free recall remained non-significant for all groups other than NHB-E, who performed significantly worse than NHW-E, and NHA-J, who performed significantly better than NHW-E (**Table S4**).

**Figure 6.**
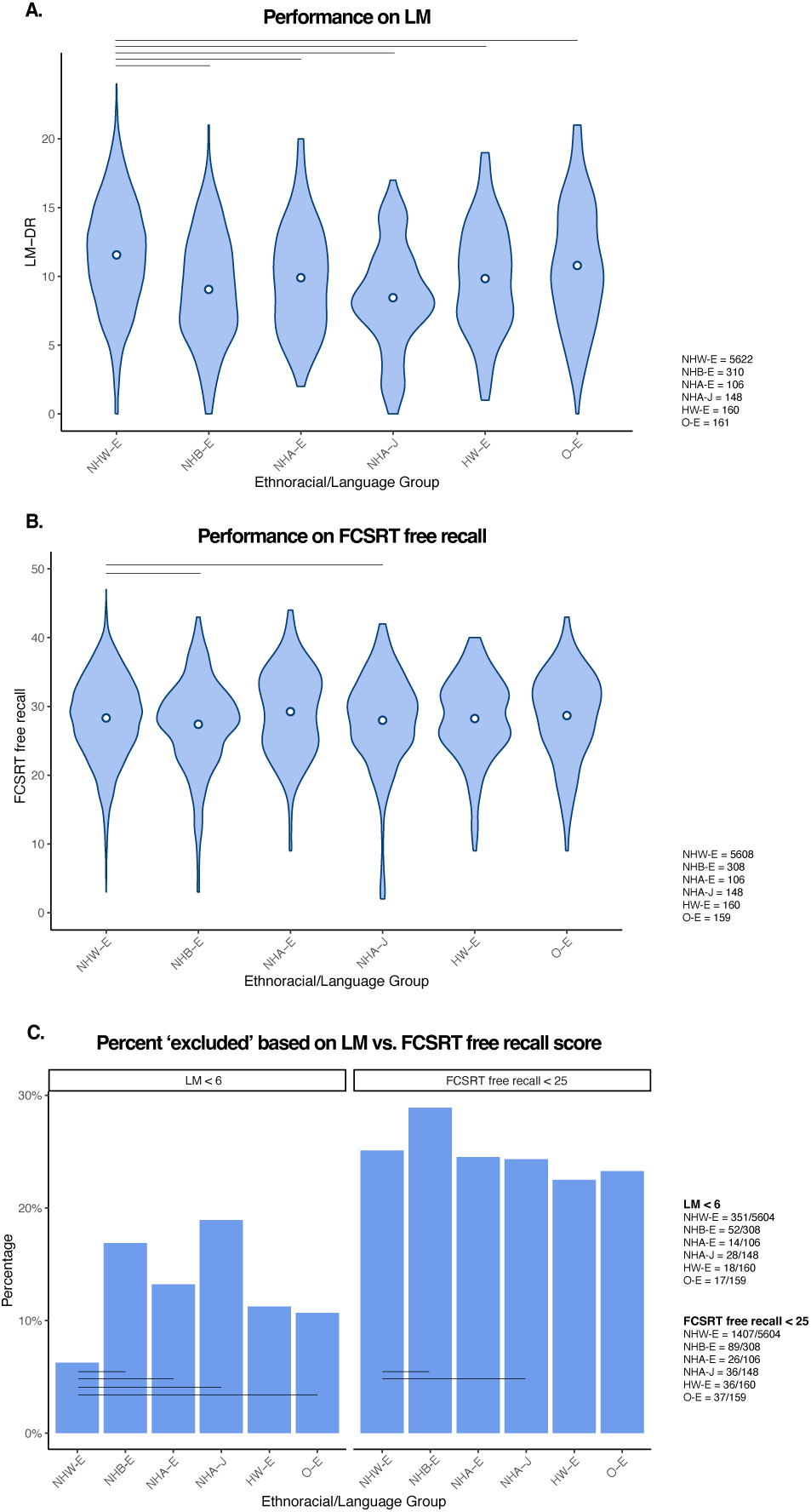
Comparison of LM and FCSRT free recall. Of all screened participants, (A) LM scores are highest in the NHW-E group compared to all other ethnoracial/language groups, whereas (B) ethnoracial/language group differences are reduced on FCSRT free recall. (C) In all screened participants who completed both LM and FCSRT, use of a FCSRT Free Recall cutoff of 25 instead of a LM cutoff of 6 would yield more balanced rates of exclusion across ethnoracial/language groups. Note: Black bars denote significant differences in LM, FCSRT free recall using linear regression and percentage of ‘excluded’ participants’ total score using logistic regression adjusting for age, sex, and education. NHW-E = Non-Hispanic White participants tested in English, NHB-E = Non-Hispanic Black participants tested in English, NHA-E = Non-Hispanic Asian participants tested in English, NHA-J = Non-Hispanic Asian participants tested in Japanese, HW-E = Hispanic White participants tested in English, O-E = Other (Unknown/Multiple Race-Ethnicities) participants tested in English. LM = Logical Memory. FCSRT = Free and Cued Selective Reminding Test.

We next examined whether using a previously established FCSRT free recall cut-off [21] instead of LM would yield a more balanced rate of exclusion across ethnoracial/language groups. If a FCSRT free recall cutoff of 25 is used instead of a LM cutoff of 6 in all participants who completed both the FCSRT and LM (n=6485), a greater percentage of participants would be excluded overall for presumed cognitive impairment (25.2% with FCSRT vs. 7.4% with LM; **Figure 6C**). However, exclusion rates would not significantly differ between ethnoracial/language groups [χ^2^(5)=3.27, p=0.66]. When accounting for age, sex, and education, with the FCSRT, NHA-E, HW-E, and O-E groups would have similar rates of ineligibility as NHW-E. Only NHB-E (28.9%) and NHA-J (24.3%) would be excluded at a significantly higher and lower rate, respectively, than NHW-E (25.8%; **Table S5**). This is in contrast to LM, for which all ethnoracial minority groups had higher rates of exclusion (11 - 19%) compared to NHW-E (6%); results are similar when accounting for age, sex, and education (**Table S5)**.

### Differing Patterns of Exclusion for Non-Cognitive Reasons

When considering all participants screened for A4 (n=6669), ethnoracial/language groups significantly differed in the percent excluded due to non-cognitive reasons (**Figure 7A)** [χ^2^(5)=27.6, p < 0.001]. Odds of exclusion due to non-cognitive reasons were significantly higher in NHB-E (24%) and O-E (25%), and significantly lower in NHA-J (10%) groups in comparison to NHW-E (17%) after accounting for age and sex (**Table S1**).

**Figure 7.**
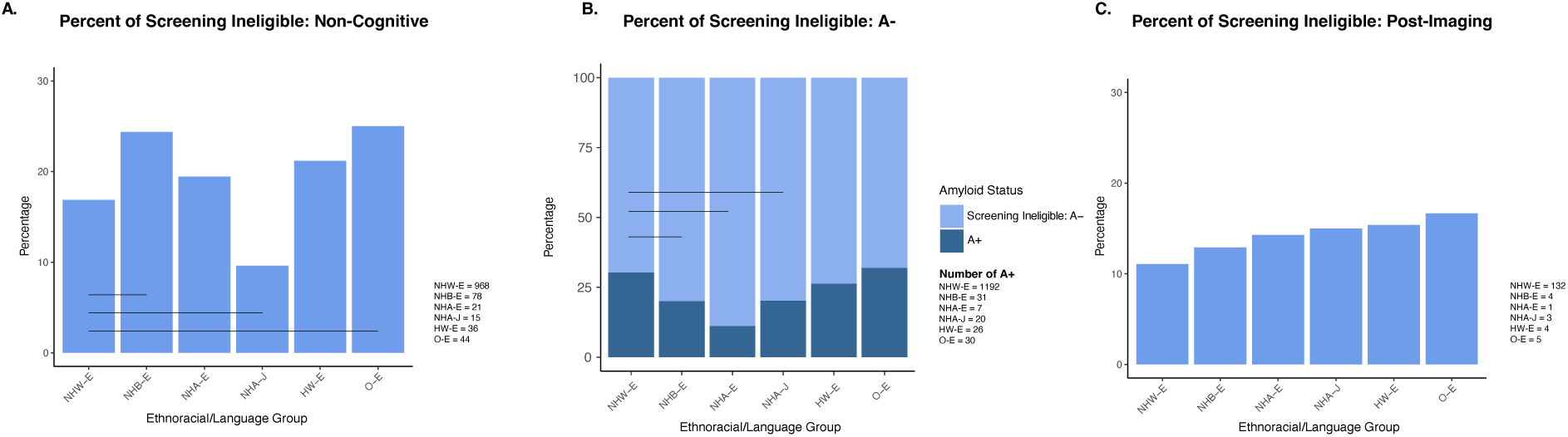
Differences in rates of screening ineligibility between ethnoracial/language group due to non-cognitive, amyloid status, and post-imaging reasons. (A) Of all screened participants, percent of exclusion due to non-cognitive reasons is higher in NHB-E and O-E, and lower in NHA-J compared to NHW-E participants. (B) In all participants who underwent amyloid PET, rate of exclusion due to A-status (i.e., lower rates of amyloid positivity) was higher in NHB-E, NHA-E, and NHA-J groups compared to NHW-E. (C) Of all A+ participants who underwent additional screening procedures after PET imaging, percent of exclusion due to post-imaging reasons were not significantly different between ethnoracial/language groups. Note: Black bars denote significant differences between groups using logistic regression adjusting for age and sex. NHW-E = Non-Hispanic White participants tested in English, NHB-E = Non-Hispanic Black participants tested in English, NHA-E = Non-Hispanic Asian participants tested in English, NHA-J = Non-Hispanic Asian participants tested in Japanese, HW-E = Hispanic White participants tested in English, O-E = Other (Unknown/Multiple Race-Ethnicities) participants tested in English. A-= Amyloid negative. A+ = Amyloid positive.

### Lower Rates of Amyloid Positivity in Ethnoracial Minorities

Of all participants who passed initial screening based on cognitive, functional, and medical criteria and thus obtained an amyloid PET scan (n=4446), 29% of participants were A+. A+ rates significantly differed between ethnoracial/language groups [χ^2^(5)=23.0, p < 0.001] (**Figure 7B**) such that A+ rates were lower in most groups compared to the NHW-E group. Significantly lower A+ rates in NHB-E, NHA-E, and NHA-J groups in comparison to NHW-E remained after accounting for age and sex (**Table S6)**.

### Exclusion Post-Imaging

When examining all participants who completed PET imaging and were A+ (n=1306), rates of post-imaging exclusion due to MRI or other reasons did not differ by ethnoracial/language group [χ^2^(5)=1.75, p=0.88] (**Figure 7C**). This pattern remained after accounting for age and sex (**Table S7**).

## DISCUSSION

In a large sample of older adults screened for a secondary prevention trial, ethnoracial minorities tested in their primary language were excluded at a higher rate than NHW-E participants because of performance on cognitive/function screening measures, non-cognitive reasons, and A-status. Of those excluded due to impaired cognition/function, the pattern of incorrect MMSE items and impaired CDR domain varied by ethnoracial/language group, indicating potential bias in test items. If FCSRT free recall was used instead of LM to exclude those with memory impairment, the overall exclusion rate would be higher but would not differ across ethnoracial/language groups. In terms of exclusion for non-cognitive reasons, NHB-E and O-E were excluded at a higher rate, and NHA-J was excluded at a lower rate than NHW-E. Together, these exclusion criteria may have downstream effects on subsequent biomarker screening as A+ rates, and thus rates of clinical trial enrollment, were lower in ethnoracial minoritized groups.

Neuropsychological tests in languages other than English are relatively limited and many tests were developed and validated in primarily NHW populations before being translated from English to other languages. There are limitations with this approach as, for example, recent examinations of Asians and Asian Americans have shown test performance differences by language, cultural context, and test version [22–24]. The MMSE, specifically, is not predictive of functional impairment in Black women [25], is affected by education and neighborhood type [26], and is impacted by testing language in bilingual participants [27]. Inclusion of the NHA-J group in A4 provides the unique opportunity to further probe the effects of language and culture on the MMSE. The NHA-J group had a unique pattern of MMSE errors with a relatively high percentage of errors on registration, some language-based (e.g., spelling WORLD backwards), and orientation-to-place items, along with a relatively low percentage of errors on recall items. Translation and/or cultural factors may have contributed to this profile. Furthermore, the MMSE has demonstrated education bias in Asian countries [28] and although the NHA-J group was fairly well-educated, their education level was still lower than all other groups.

Level of functional impairment at the CDR domain-level also differed across ethnoracial/language groups. The most drastic difference was observed in the NHA-J group, in which 100% of study partners endorsed some level of memory impairment, but did not or rarely endorsed any impairment in the domains of Home and Hobbies, Community Affairs, Personal Care, and Orientation (yet NHA-J had errors on orientation-to-place MMSE items at a higher rate than other groups). Although the sample size of NHA-J participants who completed the CDR was small (n=12), the stark difference in pattern compared to other ethnoracial/language groups is interesting and may highlight cultural differences of perceived functional impairment. Prior work has shown that CDR-SOB ratings vary by informant characteristics including relationship, gender, and frequency of contact [29], but there is no previous literature examining ethnoracial differences. Future work examining how cultural norms affect CDR ratings is needed as research on AD continues to expand globally.

LM and other story recall tasks are commonly used in research and clinical trials to evaluate memory functioning [30,31], though the exact story used may result in differing levels of bias. Although we were unable to examine individual LM items, our finding of ethnoracial/language differences on LM score is consistent with previous literature. Black older adults have consistently demonstrated lower scores on episodic memory measures, including LM, than White older adults [32–37] and this discrepancy can be explained by differences in education quality [38–41]. On the Wechsler Memory Scale, which includes LM, applying English norms to Hispanic Spanish-speaking individuals who completed translated testing in Spanish resulted in scores that were 1 standard deviation below average [42]. Although this study only investigated adults ranging from 25-34 years old, score differences may be even more exaggerated in older adults. We were unable to examine Spanish-speaking ethnoracial groups in the current study given that only 30 participants were tested in Spanish, but our results showing low LM scores in the NHA-J group may reflect a similar language-based bias. Together, these results highlight potential issues with using LM as a screening tool.

Our results show that FCSRT free recall may be a suitable alternative to LM as a screening measure to exclude those with memory impairment. However, there are important differences between FCSRT and LM to consider. First, the FCSRT includes a dedicated encoding phase that controls attention and cognitive processing, and ensures that the test stimuli are learned [43–45]. In contrast, the opportunity for learning is limited with LM as the story is only presented once. Second, the FCSRT procedure in A4 did not include a delay phase so FCSRT free recall scores reflect immediate recall [21]. In contrast, LM scores are obtained after a ∼30-minute delay [20]. Third, the FCSRT free recall score represents the total across three trials whereas there is only one recall trial for LM. Despite these differences, FCSRT free recall is strongly related to delayed recall scores on a list-learning task (B = 0.779) [46], and FCSRT free recall outperforms FCSRT total recall in predicting incident dementia [47]. A FCSRT free recall score of <25 has 90% sensitivity and 78% specificity for distinguishing between dementia and non-dementia [47], is predictive of incident dementia over 2-4 years [48], and conceptually corresponds to at least moderate retrieval impairment [21]. FCSRT free recall has been used as an episodic memory measure in AD clinical trials and has yielded similar rates of A+ and clinical progression when compared to the Repeatable Battery for the Assessment of Neuropsychological Status (RBANS), a more comprehensive screening measure of global cognition [49]. Our results showed the FCSRT free recall would exclude a greater number of participants overall than LM, but the percent of exclusion is more balanced across ethnoracial/language groups. Consistent with our finding, other studies have shown comparable cross-sectional performance between Black and White older adults in terms of free recall [50] and combined total and free recall [51], and a lack of significant race effects when predicting dementia using free recall, total recall, or combined total and free recall scores [52]. Thus, the FCSRT free recall may be a useful screening measure for clinical trials recruiting representative participants.

Our findings highlight that across both cognitive and non-cognitive screening steps, ethnoracial minority groups are excluded at higher rates compared to NHW-E participants. Biases in these procedures may then impact who obtains biomarker testing and the rate of biomarker positivity. Consistent with previous reports [14,18], we show that NHB-E, NHA-E, and NHA-J participants have lower rates of A+ than NHW-E participants in A4. Our findings demonstrate potential trickle-down effects such that biases in cognitive and non-cognitive screening procedures result in more resilient minoritized groups with less pathology. It is currently unclear if the differences in A+ rate observed here reflects clinical trial screening biases or is consistent with what is expected in the general population. In a community-based CU and mild cognitive impairment cohort, Black individuals had lower amyloid burden than NHW individuals [53,54], and in a multisite cohort study of Medicare beneficiaries with mild cognitive impairment or dementia, odds of A+ were lower for Asian, Black, and Hispanic participants in comparison to White participants [55]. However, in a review, ethnoracial differences in amyloid burden were absent when analyses were stratified by cognitive status and differences were thought to be at least partially explained by APOE-e4 status [56].

There are several limitations to consider. First, many of our groups had limited sample sizes [i.e., NHB-E (n=320), NHA-E (n=108), NHA-J (n=156), HW-E (n=170), and O-E (n=176)], especially in comparison to the NHW-E (n=5739) group. We thus focused comparisons to NHW-E, the largest A4 group and the majority of clinical trial participants broadly. Second, given the sequenced screening procedures, there was varying degrees of missingness for cognitive, functional, and medical data as all participants did not complete all assessments. Relatedly, specific reasons for medical exclusion or withdrawal were not publicly available so further patterns were not examined. Third, since the NHA-J group participated exclusively in Japan, testing language cannot be disentangled from cultural factors. Fourth, this study combined ethnic and racial categories and heterogeneity within ethnic and racial groups is important [57], but beyond the scope of this study. Finally, in A4, ethnoracial minorities were recruited through more local efforts, such as events at community churches, compared to NHW individuals, who were recruited through a larger variety of sources [14,58]. Recruitment differences may have contributed to the cognitive/function differences reported in this study.

Our study demonstrated that biases in cognitive/functional measures and initial screening procedures may have downstream effects that impact the recruitment of ethnoracial groups into clinical trials for AD. Cognitive measures that are validated for different languages and cultures and/or development of new measures that minimize biases are critically needed. Biases in screening procedures may not only disproportionately reduce the number ethnoracial minority participants who are eligible for randomization, but may also impact the characteristics of ethnoracial minorities, such as selecting particularly resilient individuals, for clinical trials.

## Data Availability

Data from the A4/LEARN study is openly available to registered researchers through https://www.a4studydata.org/.

## ACKNOWLEDGEMENTS

The A4 Study was a secondary prevention trial in preclinical Alzheimer’s disease, aiming to slow cognitive decline associated with brain amyloid accumulation in clinically normal older individuals. The A4 Study was funded by a public-private-philanthropic partnership, including funding from the National Institutes of Health-National Institute on Aging, Eli Lilly and Company, Alzheimer’s Association, Accelerating Medicines Partnership, GHR Foundation, an anonymous foundation, and additional private donors, with in-kind support from Avid Radiopharmaceuticals, Cogstate, Albert Einstein College of Medicine and the Foundation for Neurologic Diseases. The companion observational Longitudinal Evaluation of Amyloid Risk and Neurodegeneration (LEARN) Study was funded by the Alzheimer’s Association and GHR Foundation. The A4 and LEARN Studies were led by Dr. Reisa Sperling at Brigham and Women’s Hospital, Harvard Medical School, and Dr. Paul Aisen at the Alzheimer’s Therapeutic Research Institute (ATRI) at the University of Southern California. The A4 and LEARN Studies were coordinated by ATRI at the University of Southern California, and the data are made available under the auspices of Alzheimer’s Clinical Trial Consortium through the Global Research & Imaging Platform (GRIP). The complete A4 Study Team list is available on. We would like to acknowledge the dedication of the study participants and their study partners who made the A4 and LEARN Studies possible.

## CONSENT STATEMENT

The current study did not access identifying information and are therefore not considered human subjects research.

