## Supplementary Tables for "Cognitive screening biases in a secondary prevention Alzheimer’s disease clinical trial"

[Table S5. Odds of “exclusion” due to LM < 6 vs. FCSRT free recall < 25 across ethnoracial/language groups out of all participants who completed both tests.	6](#_Toc215058742)

**Table S1. Odds of exclusion due to cognitive/functional and non-cognitive reasons across ethnoracial/language groups out of all screened participants.** The logistic regression model included fixed effects of age (centered at 72 years), sex, ethnoracial/language group, and education (centered at 16 years) for cognitive screening ineligibility. Unstandardized beta estimates, standard errors, and p values are reported. Bolded cells indicate a significant difference from non-Hispanic White participants tested in English (NHW-E).

Note: NHB-E = Non-Hispanic Black participants tested in English, NHA-E = Non-Hispanic Asian participants tested in English, NHA-J = Non-Hispanic Asian participants tested in Japanese, HW-E = Hispanic White participants tested in English, O-E = Other (Unknown/Multiple Race-Ethnicities) participants tested in English.

|  | Odds of Screening Ineligible:  Cognition/Function | Odds of Screening Ineligible:  Non-Cognitive |
| --- | --- | --- |
| NHB-E vs.  NHW-E | **0.767 (0.135), <0.001** | **0.478 (0.135),**  **<0.001** |
| NHA-E vs.  NHW-E | **0.569 (0.237), 0.017** | 0.155 (0.246),  0.528 |
| NHA-J vs.  NHW-E | **0.550 (0.187), 0.003** | **-0.680** (**0.274),**  **0.013** |
| HW-E vs.  NHW-E | 0.351 (0.198), 0.076 | 0.274 (0.191),  0.152 |
| O-E vs.  NHW-E | **0.493 (0.199), 0.013** | **0.509 (0.178),**  **0.004** |

**Table S2. Odds of exclusion due to impaired cognition/functioning across ethnoracial/language groups out of all screened participants.** The logistic regression model included fixed effects of age (centered at 72 years), sex, education (centered at 16 years), and ethnoracial/language group. Unstandardized beta estimates, standard errors, and p values are reported. Bolded cells indicate a significant difference from non-Hispanic White participants tested in English (NHW-E).

Note: NHB-E = Non-Hispanic Black participants tested in English, NHA-E = Non-Hispanic Asian participants tested in English, NHA-J = Non-Hispanic Asian participants tested in Japanese, HW-E = Hispanic White participants tested in English, O-E = Other (Unknown/Multiple Race-Ethnicities) participants tested in English.

|  | Odds of Screening Ineligible: Impaired Cognition/Function |
| --- | --- |
| NHB-E vs. NHW-E | **1.035 (0.140), <0.001** |
| NHA-E vs. NHW-E | **0.746 (0.253), 0.003** |
| NHA-J vs. NHW-E | **0.756 (0.190), <0.001** |
| HW-E vs. NHW-E | **0.509 (0.209), 0.015** |
| O-E vs. NHW-E | **0.617 (0.214), 0.004** |

**Table S3. Performance on MMSE and CDR-SOB across ethnoracial/language groups out of all screened participants.** The linear regression model included fixed effects of age (centered at 72 years), sex, education (centered at 16 years), and ethnoracial/language group. Unstandardized beta estimates, standard errors, and p values are reported. Bolded cells indicate a significant difference from non-Hispanic White participants tested in English (NHW-E).

Note: NHB-E = Non-Hispanic Black participants tested in English, NHA-E = Non-Hispanic Asian participants tested in English, NHA-J = Non-Hispanic Asian participants tested in Japanese, HW-E = Hispanic White participants tested in English, O-E = Other (Unknown/Multiple Race-Ethnicities) participants tested in English.

|  | MMSE (N=6482) | CDR-SOB (N=6023) |
| --- | --- | --- |
| NHB-E vs. NHW-E | **-0.711 (0.085), <0.001** | **0.169 (0.024),**  **<0.001** |
| NHA-E vs. NHW-E | **-0.707 (0.142), <0.001** | **0.142 (0.039),**  **<0.001** |
| NHA-J vs. NHW-E | **-0.356 (0.121), 0.003** | 0.0315 (0.035),  0.384 |
| HW-E vs. NHW-E | **-0.634 (0.116),** **<0.001** | **0.130 (0.032),**  **<0.001** |
| O-E vs. NHW-E | **-0.334 (0.119), 0.005** | **0.090 (0.033),**  **0.006** |

**Table S4. Performance on LM and FCSRT free recall across ethnoracial/language groups out of all screened participants.** The linear regression model included fixed effects of age (centered at 72 years), sex, education (centered at 16 years), and ethnoracial/language group. Unstandardized beta estimates, standard errors, and p values are reported. Bolded cells indicate a significant difference from non-Hispanic White participants tested in English (NHW-E).

Note: NHB-E = Non-Hispanic Black participants tested in English, NHA-E = Non-Hispanic Asian participants tested in English, NHA-J = Non-Hispanic Asian participants tested in Japanese, HW-E = Hispanic White participants tested in English, O-E = Other (Unknown/Multiple Race-Ethnicities) participants tested in English.

|  | LM (N=6507) | FCSRT free recall (N=6489) |
| --- | --- | --- |
| NHB-E vs. NHW-E | **-2.309 (0.230), <0.001** | **-1.167 (0.338),**  **0.001** |
| NHA-E vs. NHW-E | **-1.719 (0.383), <0.001** | 0.987 (0.561),  0.079 |
| NHA-J vs. NHW-E | **-2.435 (0.328), <0.001** | **1.018 (0.481),**  **0.033** |
| HW-E vs. NHW-E | **-1.402 (0.315),** **<0.001** | 0.120 (0.462),  0.671 |
| O-E vs. NHW-E | **-0.829 (0.319), 0.009** | 0.122 (0.470),  0.800 |

**Table S5. Odds of “exclusion” due to LM < 6 vs. FCSRT free recall < 25 across ethnoracial/language groups out of all participants who completed both tests**. Logistic regression with fixed effects of age (centered at 72 years), sex, education (centered at 16 years) and ethnoracial/language group. Unstandardized beta estimates, standard errors, and p values are reported. Bolded cells indicate a significant difference from non-Hispanic White participants tested in English (NHW-E).

Note: NHB-E = Non-Hispanic Black participants tested in English, NHA-E = Non-Hispanic Asian participants tested in English, NHA-J = Non-Hispanic Asian participants tested in Japanese, HW-E = Hispanic White participants tested in English, O-E = Other (Unknown/Multiple Race-Ethnicities) participants tested in English. LM = Logical Memory. FCSRT = Free and Cued Selective Reminding Test.

|  | LM < 6 | FCSRT free recall < 25 |
| --- | --- | --- |
| NHB-E vs. NHW-E | **1.053 (0.169), <0.001** | **0.289 (0.138), 0.036** |
| NHA-E vs. NHW-E | **0.924 (0.300), 0.002** | -0.051 (0.238), 0.830 |
| NHA-J vs. NHW-E | **0.935 (0.223), <0.001** | **-0.431 (0.201), 0.032** |
| HW-E vs. NHW-E | 0.492 (0.267), 0.066 | -0.233 (0.202), 0.249 |
| O-E vs. NHW-E | **0.601 (0.276), 0.029** | -0.055 (0.203), 0.788 |

**Table S6. Odds of amyloid positivity at PET across ethnoracial/language groups out of all participants who completed amyloid PET.** Logistic regression with fixed effects of age (centered at 72 years), sex, and ethnoracial/language group. Unstandardized beta estimates, standard errors, and p values are reported. Bolded cells indicate a significant difference from non-Hispanic White participants tested in English (NHW-E).

Note: NHB-E = Non-Hispanic Black participants tested in English, NHA-E = Non-Hispanic Asian participants tested in English, NHA-J = Non-Hispanic Asian participants tested in Japanese, HW-E = Hispanic White participants tested in English, O-E = Other (Unknown/Multiple Race-Ethnicities) participants tested in English.

|  | Odds of A+ |
| --- | --- |
| NHB-E vs. NHW-E | **-0.539 (0.205), 0.009** |
| NHA-E vs. NHW-E | **-1.244 (0.403), 0.002** |
| NHA-J vs. NHW-E | **-0.582 (0.255), 0.022** |
| HW-E vs. NHW-E | -0.232 (0.232), 0.319 |
| O-E vs. NHW-E | 0.092 (0.225), 0.682 |

**Table S7. Odds of exclusion due to post-imaging reasons across ethnoracial/language groups out of all A+ participants after amyloid PET.** Logistic regression with fixed effects of age (centered at 72 years), sex, and ethnoracial/language group. Unstandardized beta estimates, standard errors, and p values are reported.

Note: NHW-E = non-Hispanic White participants tested in English, NHB-E = Non-Hispanic Black participants tested in English, NHA-E = Non-Hispanic Asian participants tested in English, NHA-J = Non-Hispanic Asian participants tested in Japanese, HW-E = Hispanic White participants tested in English, O-E = Other (Unknown/Multiple Race-Ethnicities) participants tested in English.

|  | Odds of Screening Ineligible:  Post-Imaging |
| --- | --- |
| NHB-E vs. NHW-E | 0.148 (0.547), 0.787 |
| NHA-E vs. NHW-E | 0.385 (1.089), 0.724 |
| NHA-J vs. NHW-E | 0.097 (0.642), 0.879 |
| HW-E vs. NHW-E | 0.284 (0.556), 0.609 |
| O-E vs. NHW-E | 0.487 (0.502), 0.332 |
